# Wearable Electrodermal Activity Reveals Sustained Sympathetic Responses to Sleep Arousals

**DOI:** 10.64898/2026.02.19.26346633

**Authors:** Tugce Canbaz Gumussu, Hugo F. Posada-Quintero, Youngsun Kong, Camila Jimenez Wong, Ki H. Chon, Walter Karlen

## Abstract

**Introduction:** Sympathetic responses to sleep arousals are well documented through cardiac measures, yet capturing autonomic physiology across additional channels offers a more complete picture of the body’s reaction to these brief events. Electrodermal activity (EDA), driven by sympathetic sudomotor pathways, offers a distinct and largely unexplored channel to characterize autonomic responses during sleep arousals.

**Methods:** To characterize autonomic reactivity during sleep arousals, we analyzed synchronous polysomnography and wrist-worn EDA recordings from 100 adults of the DREAMT dataset. We applied a time-varying frequency decomposition framework to isolate sleep-specific sympathetic components, extracting statistical and peak-based features from arousal events and matched stable-sleep controls. By concurrently assessing heart rate, we captured a multimodal profile of the autonomic arousal response. Finally, we evaluated the modulatory effects of arousal duration and sleep stage on EDA, alongside the confounding influence of wrist movement.

**Results:** Relative to stable-sleep controls, arousals elicited a robust, sustained sympathetic EDA modulation that persisted up to 40 s post-arousal. In contrast, heart rate exhibited a sharp, transient increase that rapidly returned to baseline following the event. We observed that arousal duration significantly modulated sudomotor reactivity, with long arousals generating prominent responses compared to the weaker activity elicited by short arousals. Furthermore, while EDA feature trajectories remained consistent across REM and NREM sleep, REM periods introduced greater signal variability. Crucially, wrist movement did not fundamentally drive the EDA response as sympathetic activation persisted during minimal-movement arousals, though concurrent movement amplified the signal. Ultimately, our analysis revealed that this prolonged EDA activation is primarily driven by clustered sympathetic bursts and amplitude enhancement, rather than shifts in peak frequency.

**Conclusion:** Our findings underscore the value of multimodal perspective, demonstrating that wearable EDA captures autonomic information distinct from conventional cardiac measures. Together, these signals establish a comprehensive, quantitative baseline for tracking sympathetic dynamics during sleep arousals.

## 1 INTRODUCTION

Sleep arousals are characterized by short altered brain states that often evoke transient autonomic activations (Halász et al., 2004). Prior work using cardiac markers such as heart rate (Sforza et al., 2004) and heart rate variability (Blasi et al., 2003) has suggested that these autonomic responses are associated with increased sympathetic nervous system (SNS) activity. The sympathetic nervous system governs a wide range of peripheral responses beyond cardiac regulation, including sudomotor activity or microneurography (de Zambotti et al., 2018). Although autonomic responses with the cardiac measures are well described, characterization of arousal-related autonomic physiology through different sympathetic channels offers a more complete picture of how the body responds to these brief events.

Electrodermal activity (EDA) is an emerging signal in wearable sensing, driven exclusively by sympathetic sudomotor pathways (Boucsein, 2012), making it a specific peripheral marker of sympathetic activity. The eccrine sweat glands in the skin are exclusively innervated by sympathetic nerve fibers (Cui and Schlessinger, 2015), and their activation increases skin moisture, thereby enhancing electrical conductivity. Given that cardiac studies point to sympathetic activation during sleep arousals, EDA provides a complementary modality for investigating sympathetic engagement more specifically. As EDA can be measured continuously using compact wearable sensors, understanding its behavior during sleep arousals may facilitate future unobtrusive monitoring of sleep-related autonomic function.

While EDA has long been used to investigate physiological arousal during wakefulness (Sánchez-Reolid et al., 2022), its application to sleep has gained increasing attention only in recent years. Previous studies have characterized EDA activity across sleep stages (Sano et al., 2014) and during deep sleep (Onton et al., 2016), and have explored its utility for sleep staging (Anusha et al., 2022) and sleep–wake classification (Herlan et al., 2019). Despite these advances, little is known about how EDA behaves during sleep arousals. In particular, no prior work has systematically characterized the temporal dynamics of EDA across arousal events or compared arousal-related EDA responses with stable, non-arousal sleep periods.

In this work, we investigate physiological responses during sleep arousals using EDA and characterize how EDA-derived features evolve from before to well after arousal onset (Figure 1). We further compare these responses with heart rate, a conventional autonomic marker, and investigate how movement intensity relates to the observed sympathetic response patterns. Our main contributions are as follows:

- We introduce a frequency-domain characterization of EDA during sleep to identify the spectral components most dominant during stable sleep and arousals.
- We provide the first systematic, segment-wise analysis of EDA dynamics across arousal events, revealing arousal-specific sympathetic responses distinct from those during stable sleep.
- We compare EDA-derived measures with heart rate to investigate the complementary physiological information provided by the two modalities during sleep arousals.
- We characterize the relationship between physical movement intensity and the observed EDA response patterns during arousals.

**Figure 1.**
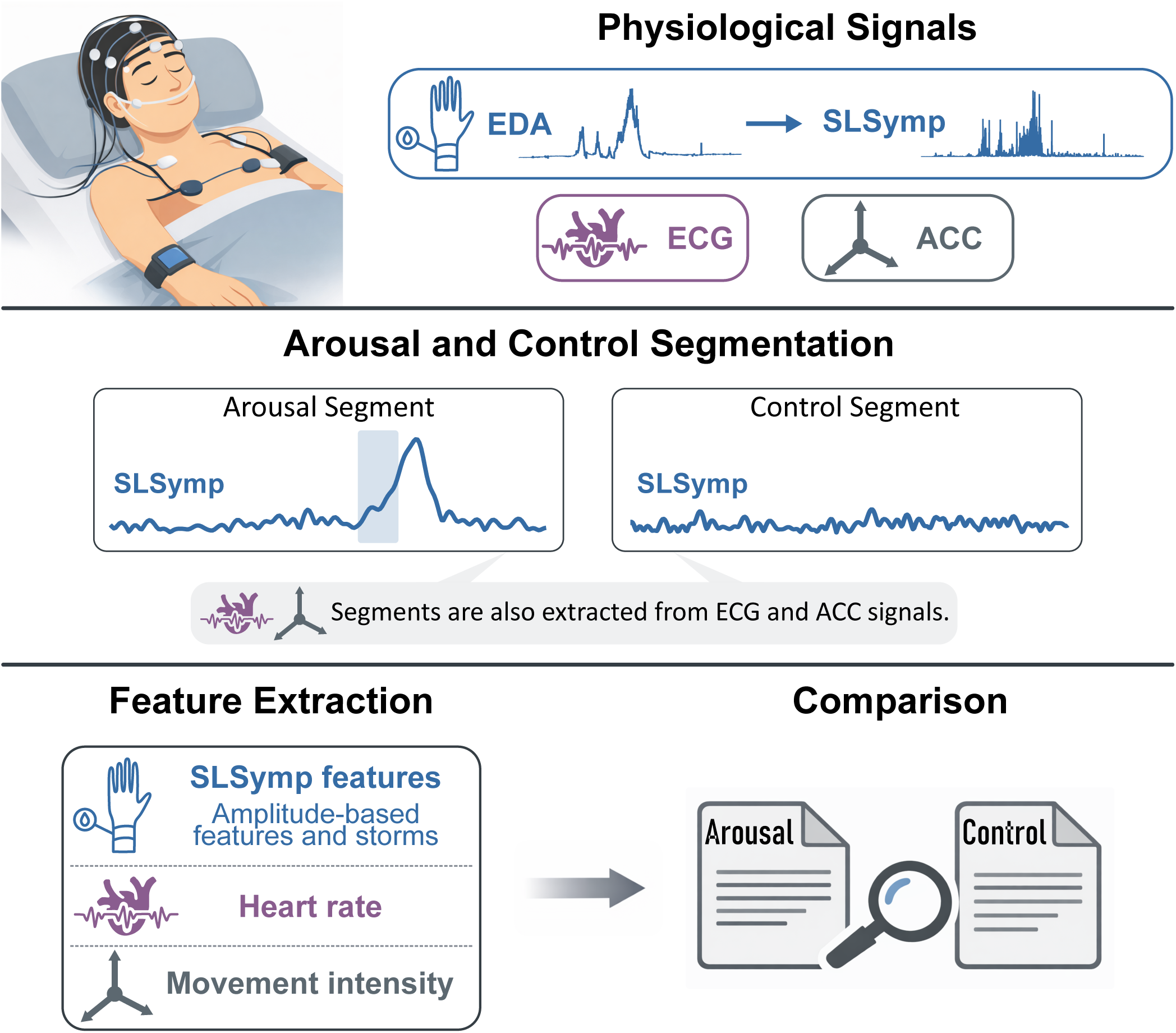
Overview of the multimodal analysis during sleep arousals. We derived a sleep-specific sympathetic component (SLSymp) from wrist electrodermal activity (EDA), and used electrocardiogram (ECG) and wrist accelerometer (ACC) as complementary signals. From each signal, we extracted arousal and matched control segments across all three modalities. We computed features from SLSymp, ECG, and ACC to compare arousal and control segments and characterize arousal-related physiological dynamics.

## 2 METHODS

Our aim was to characterize physiologic responses associated with sleep arousals using EDA, electrocardiogram (ECG) and accelerometer (ACC). We combined time-frequency decomposition with structured signal segmentation to track how the EDA-derived sympathetic response evolves before and after arousal onset. By comparing arousal-centered segments with duration-matched control segments from stable sleep, we isolated arousal-specific dynamics from the physiological variability naturally present during stable sleep, and further assessed how arousal duration and sleep stage influenced these responses. To contextualize the EDA-derived responses, we extracted heart rate from the ECG as a conventional cardiac marker of arousal, and examined wrist movement from the ACC to assess whether the observed sympathetic dynamics could be explained by motion rather than genuine autonomic activation. We analyzed an open-source dataset with wrist-recorded EDA and accelerometer signals and polysomnography-derived ECG, obtaining a systematic evaluation of sympathetic activation during sleep arousals.

### 2.1 EDA Pre-Processing

Sympathetic activity fluctuates continuously and prior work has shown that time-varying analyses of EDA can provide sensitive markers of physiological arousal (Posada-Quintero et al., 2016, 2018). To capture these fluctuations during sleep, we used a frequency-selective, time-varying decomposition approach.

We used variable-frequency complex demodulation (VFCDM) (Wang et al., 2006) to decompose the EDA signal into narrowband oscillatory components, similar to previously proposed frequency-domain characterizations of EDA (Posada-Quintero et al., 2016). We used VFCDM because it provides higher time-frequency resolution and more reliable amplitude estimation than alternative approaches, such as wavelet transform (Wang et al., 2006). After decomposition, to obtain the instantaneous amplitude, we applied the Hilbert transform to the sum of the reconstructed components. Only components within the activity-specific frequency bands were reconstructed.

#### 2.1.1 Determining Dominant EDA Frequency Bands

As the dominant frequency range for sleep-related sympathetic responses has not been firmly established, we first performed a frequency-domain characterization to guide band selection. We analyzed two types of 1-min EDA windows: (1) *reference windows*, drawn from stable sleep without arousals, apneas, or wake epochs; and (2) *arousal-onset windows*, starting at arousal onset but still free of apnea or wake contamination. These windows allowed us to compare baseline spectral content with arousal-associated spectral changes. We high-pass filtered (fourth-order Butterworth, 0.01 Hz cutoff) the 4 Hz EDA to remove baseline drift and its harmonics. For each window, we estimated the power spectral density using Welch’s method (60-sample windows, 50% overlap). We identified dominant frequency bands among eight low-frequency EDA bands (Table 1). For each participant, we computed the relative power of each band *P*_b_ as a percentage of total power 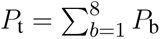, yielding a percentage contribution (*P*_b_*/P*_t_) × 100 for each band. We then sorted these percentage values in descending order to obtain a ranked power profile. To identify the change point in this profile, we computed the slope between each pair of consecutive ranked power values and took the ratio of each slope to the next. The maximum slope ratio indicated the point where the largest relative deceleration in the power decline occurs, marking the transition from steep to comparatively shallow. We defined the bands ranked above this transition point as dominant, as they collectively account for the most rapid accumulation of power before the profile flattens.

**Table 1.**
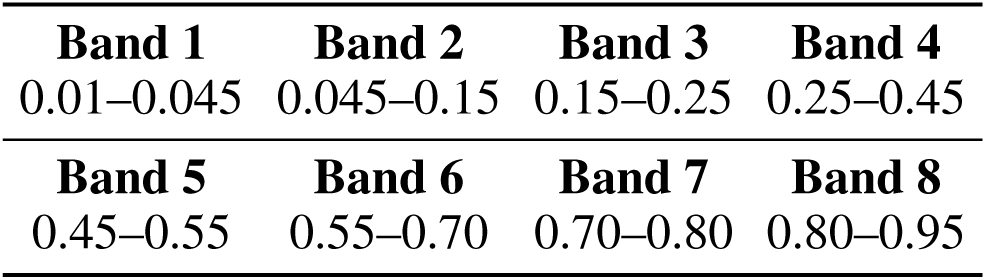
Frequency bands (Hz) used for EDA decomposition.

#### 2.1.2 Extracting the Sleep-Specific Sympathetic Component - SLSymp

After extracting dominant EDA frequencies during sleep, we extracted the sleep-specific sympathetic component from the raw EDA signal using the dominant bands. We high-pass filtered the raw EDA signal using a fourth-order Butterworth filter (0.01 Hz cutoff) to remove tonic drifts. We then extracted sympathetic tone by applying VFCDM restricted to the selected bands. For this, let *x*(*t*) denote the high-pass filtered EDA signal. VFCDM reconstructs the selected components, whose sum we denote as *X*′(*t*). We normalize *X*′(*t*) by its standard deviation, compute its Hilbert transform and obtain *Y*′(*t*). Together, *X*′(*t*) and *Y*′(*t*) form a complex conjugate pair that defines the analytic signal *Z*(*t*) as

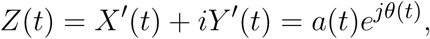

where

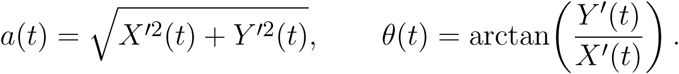

We defined the instantaneous amplitude *a*(*t*) as SLSymp, denoting its role as a sleep-specific index of sympathetic activation.

To assess the contribution of the selected frequency bands, we additionally computed a complementary signal using only the excluded bands. We performed all subsequent analyses on the SLSymp signal derived from the dominant bands to characterize how sympathetic activity evolves around sleep arousal events.

### 2.2 ECG Pre-Processing

ECG is a commonly used source of investigating autonomic activity during sleep (Sforza et al., 2004), and we included it as a reference against which the EDA-derived sympathetic responses can be evaluated. To prepare ECG for feature extraction, we applied a third-order Butterworth bandpass filter (1-40 Hz) to remove baseline wander and high-frequency noise, and standardized the filtered signal to zero mean and unit variance.

### 2.3 ACC Pre-Processing

As body movement can co-occur with, and potentially influence, EDA responses, we included the triaxial wrist ACC signal in our analyses. We computed the Euclidean norm across the three ACC axes and removed the gravitational bias using an exponentially weighted moving average high-pass filter (*α* = 0.1). We then applied a two-sample causal moving average to smooth transient fluctuations and took the absolute value of the resulting ACC signal.

### 2.4 Segment Extraction

To analyze how sympathetic activity evolves around sleep arousal events, we divided the SLSymp and ECG into arousal and matched control segments. For each annotated arousal, we extracted a segment of approximately 80 s, with exact duration depending on the length of the arousal. For every arousal segment, we also generated a corresponding control segment matched in structure and duration from stable sleep without arousals. We divided all arousal and control segments into consecutive 10 s windows, which served as the units of analysis. The 10 s window length was chosen because electrodermal responses evolve slowly, and this duration provides sufficient temporal resolution while still capturing meaningful sympathetic activity changes.

#### Arousal Segments

For each labeled arousal, we extracted a segment beginning 10 s before arousal onset and extending 60 s beyond arousal end. Because arousal durations varied, segments exceeded 80 s whenever the arousal lasted longer than 10 s. For arousals shorter than 10 s, we imposed a minimum duration of 10 s (i.e., arousal end = onset + 10 s) to ensure a meaningful analysis window.

Each arousal segment was partitioned into consecutive 10 s windows without overlap (Figure 2):

- **W-1:** the 10 s window preceding arousal onset,
- **W0:** the window covering the entire arousal duration (minimum 10 s),
- **W1–W6:** six consecutive 10 s windows following the W0 window.

**Figure 2.**
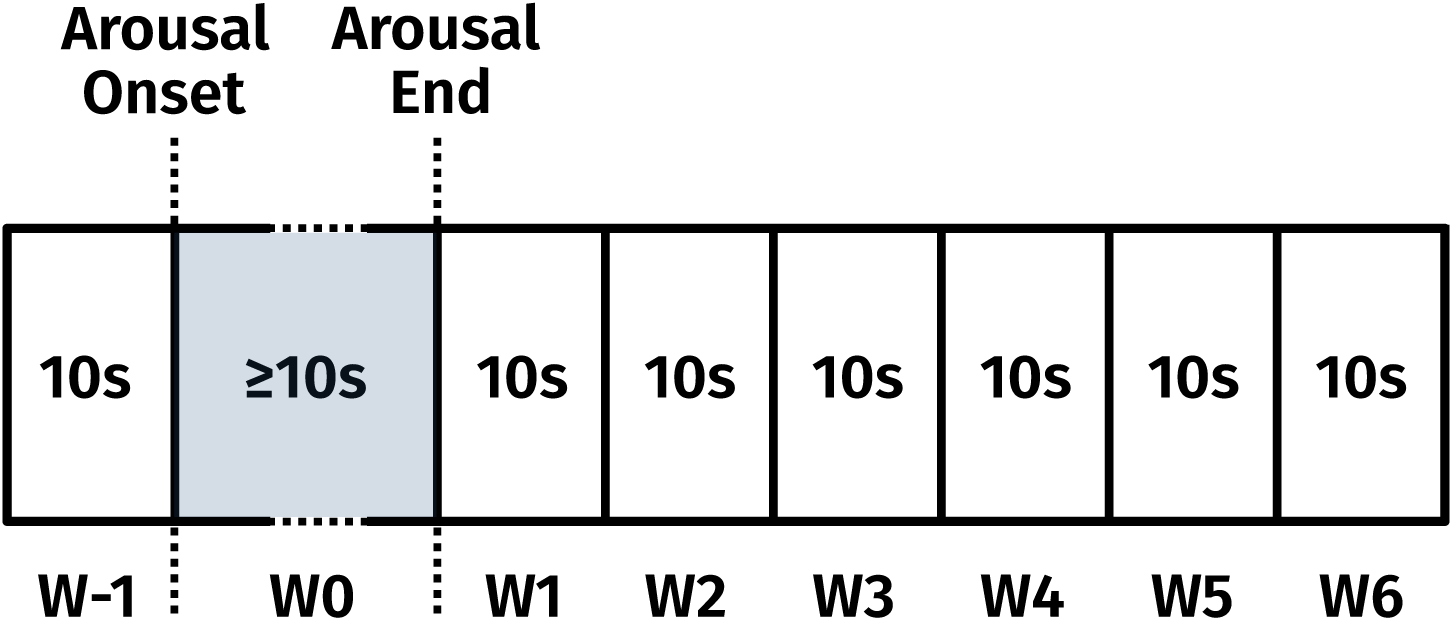
Windowing of arousal segments. W-1 is the 10 s window before arousal onset. W0 starts with the arousal onset and includes the entire arousal. The minimum duration of W0 window is 10 s. The windows from W1 to W6 are consecutive 10 s windows after W0.

We included only arousals occurring during sleep. Any segment containing wake epochs within its windows was discarded. Because apneic events substantially influence sympathetic activity (Bisogni et al., 2016; Dissanayake et al., 2021), we excluded arousals associated with apnea. An arousal was considered apnea-associated if either its W-1 or W0 window overlapped with an apneic event, consistent with the physiological pattern in which apneas commonly terminate with an arousal and resumption of breathing (Huang et al., 2022).

#### Control Segments

For each arousal, we randomly selected a matched control segment from stable sleep that was free of arousals, apneas, and wake epochs. Each control segment mirrored its corresponding arousal segment in total duration, number of windows, sleep stage and window lengths (W-1, W0, and W1–W6). This one-to-one matching allowed direct, window-wise comparisons between arousal-related and stable sleep dynamics while controlling for segment structure and temporal context.

### 2.5 Feature Extraction

For each 10 s window (W-1, W0, W1–W6) in both arousal and matched control segments, we extracted features from the SLSymp and the ECG to quantify sympathetic dynamics across the arousal timeline.

#### 2.5.1 EDA Features

The four amplitude-based features can be listed as:

- **Min amplitude:** minimum SLSymp value within the window,
- **Max amplitude:** maximum SLSymp value within the window,
- **Min–max amplitude:** difference between maximum and minimum SLSymp values,
- **Standard deviation (SD):** sample standard deviation of SLSymp values.

In addition to amplitude-based features, we quantified peak dynamics across the (≥70 s) interval spanning the W0 through W6 windows. Prior work introduced *storm*, defined as repeated EDA peaks occurring within a short time window during sleep (Sano et al., 2014; Burch, 1965). We classified a segment as a storm if it contained a minimum number of peaks within the analysis window. To evaluate whether arousals not only increase the likelihood of peak occurrence but also affect the number of peaks, we examined storms defined by at least two, three, and four peaks. For each participant, we detected storms and computed their proportion relative to the total number of available segments. Because no consensus exists regarding the optimal threshold for defining a peak, we detected storms across multiple peak amplitude thresholds (0.05, 0.1, 0.2, 0.3, and 0.4 µS).

We focused our analysis on SLSymp, which captures the fast components of EDA signals and characterizes sympathetic dynamics. We did not include skin conductance level (SCL) features as SCL reflects slow tonic changes in EDA that can be influenced by many confounding factors, such as hydration level, making it less optimal for our analysis. We also excluded conventional skin conductance response (SCR) features, which quantify rapid transient EDA changes in the time domain (e.g., SCR rate or amplitude). Prior work has shown that time-varying spectral analyses of EDA capture information comparable to these SCR measures (Posada-Quintero and Chon, 2020).

#### 2.5.2 Heart Rate

Heart rate (HR) is a well-established autonomic marker of arousal during sleep and has been widely used to characterize the cardiac component of the arousal response (Sforza et al., 2004). We included HR in our analysis to compare the dynamics of EDA-derived sympathetic activity against a conventional sympathetic measure.

We first detected R-peaks in the filtered ECG signal using the Pan-Tompkins approach (Pan and Tompkins, 1985). We applied a derivative filter to enhance the QRS complex, squared the output to amplify peaks, and smoothed the result with a 150 ms moving average window. We detected R-peaks with a minimum inter-peak distance of 300 ms, corresponding to a maximum physiological rate of 200 beats-per-minute (bpm), and a minimum peak height of 30% of the signal maximum. We computed R-R intervals from consecutive R-peak locations and rejected intervals outside the physiologically plausible range of 30-200 bpm. We then converted the remaining R-R intervals to instantaneous HR values in bpm.

#### 2.5.3 Movement Intensity

For each arousal and control segment, we extracted movement intensity across all windows (W-1 to W6) to characterize the degree to which arousals are accompanied by physical movement relative to stable sleep. The movement intensity corresponded to the peak amplitude of the preprocessed ACC signal within each window. We classified a window as high activity if its movement intensity exceeded the mean peak amplitude of control segments, and as low activity otherwise.

### 2.6 Event Analysis

We excluded segments with high wrist activity in W-1, as this could interfere with SLSymp changes in the following windows. For each feature, we fitted a linear mixed-effects model (LMM) with window (W-1, W0, W1–W6), condition (arousal, control), and their interaction as fixed effects, and subject as a random intercept.

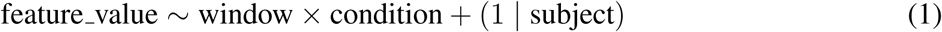

We applied a type III analysis of variance (ANOVA) to this model to test the main effects of window and condition and their interaction. A significant *window×condition* interaction (*p <* 0.05) indicates that the temporal trajectory across windows differs between arousal and control segments. When this interaction was significant, we used estimated marginal means (EMM) with Holm-adjusted pairwise comparisons to test window-to-window differences within each condition, focusing on comparisons of each window against W-1 to characterize when each condition’s response became significantly different from baseline. We applied this same model structure separately to REM and NREM arousal and control segments to assess whether arousal-related feature changes reflect general autonomic activation or stage-dependent mechanisms.

Because arousal duration modulates autonomic responses (Azarbarzin et al., 2014), we investigated if the arousal length influences SLSymp changes. For this, we treated duration as a continuous predictor in a LMM. Arousal duration was z-scored prior to modeling to improve numerical stability. We fitted the model:

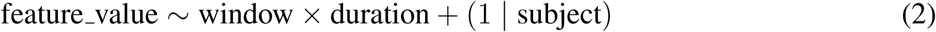

where *feature_value* is the values of the SLSymp features, *window* is the categorical time window factor with W-1 as the reference level, *duration* is the z-scored arousal duration, and subject is a random intercept to account for repeated measurements within individuals. The interaction term *window×duration* tested whether the association between arousal duration and SLSymp feature magnitude differs across post-arousal windows. We assessed the significance of fixed-effect interaction coefficients using Satterthwaite-approximated degrees of freedom, and corrected *p*-values for multiple comparisons across windows using the Holm method. To visualize this pattern, we grouped arousals into three duration categories using the 25th and 75th percentiles of the duration distribution as cutoffs, and plotted the mean SLSymp feature trajectory within each group.

We visually inspected Q-Q plots of the model residuals to confirm the assumption of normality. Additionally, we checked all model fits to ensure absence of singularity.

For the visual comparison across segments, we expressed each feature trajectory as change from the pre-arousal baseline in the figures. Specifically, we subtracted each segment’s W-1 value from all subsequent windows, so that all trajectories start at zero and reflect the magnitude of change from the pre-arousal level (ΔW).

In addition to the amplitude-based feature analysis, we compared the proportion of storms between arousal and control segments across all amplitude thresholds. This threshold-resolved comparison allowed us to evaluate whether differences in storm occurrence remained consistent regardless of the specific peak-detection criterion.

### 2.7 Movement Analysis

Body movement during sleep is associated with sympathetic activity Senel and DelRosso (2026), and as a result, physical movement and sympathetic activation can naturally co-occur during arousals. In this analysis, we aimed to characterize how the SLSymp response evolved under different levels of wrist movement, and to describe the response pattern even when movement was minimal. Unlike Section 2.6, we did not exclude high-activity segments in W-1 here, as our goal was to characterize the movement–SLSymp relationship rather than control for it.

We examined how the timing of the wrist movement related to the observed SLSymp trends. We grouped arousals by the window in which high activity was detected and computed the mean SLSymp (max amplitude feature, ΔW) trajectory across all windows for each group. We also computed the mean SLSymp trajectory for arousals with low activity across all analyzed windows. This allowed us to characterize how SLSymp evolved under high and low levels of movement, and to assess whether the observed EDA response patterns persisted even when the wrist movement was minimal. For each group separately, we applied the same LMM framework described for arousal duration analysis, fitting *feature_value ~ window* + (1 | *subject*) with W-1 as the reference level to test which post-arousal windows showed significant change from baseline within each movement group.

### 2.8 Dataset

We analyzed the open-source DREAMT Dataset for Real-time Sleep Stage Estimation using Multisensor Wearable Technology (Wang et al., 2025, 2024), available on PhysioNet (Goldberger et al., 2000). The dataset contained recordings from 100 participants, including both individuals without sleep disorders and those with sleep-related conditions such as obstructive sleep apnea, excessive daytime sleepiness, or restless leg syndrome (RLS). We excluded the participants with RLS because the associated leg movements were not annotated in the dataset and could interfere with our arousal analysis. In addition, we excluded recordings with constant EDA channels, as they do not contain meaningful EDA dynamics.

Overnight polysomnography (PSG) recordings included electroencephalography (EEG), electromyography (EMG), electrooculography, ECG, respiratory effort, airflow, and oxygen saturation signals sampled at 200 Hz with a Nihon Kohden Polysmith 1004 (version 11) data management system. Additionally, the participants had worn the Empatica E4 wristband (Empatica Inc., Milano, Italy; software version Summer 2019) on the left wrist, which provided photoplethysmography sampled at 64 Hz, ACC at 32 Hz, and temperature and EDA at 4 Hz. All wearable and PSG signals had been synchronized by timestamp alignment and resampled to 100 Hz to ensure a unified sampling rate across modalities. Sleep stages (Wake, N1, N2, N3) had been manually scored in 30 s epochs by sleep technicians, with annotations available for sleep apnea events and RLS diagnosis.

For our analysis, we downsampled the EDA signal from 100 Hz to 4 Hz after applying an anti-aliasing filter (fourth-order Butterworth, 2 Hz cutoff). We used ECG and ACC at 100 Hz without resampling.

### 2.9 EDA Signal Quality Analysis

We assessed EDA signal quality using an open-source pre-processing toolbox for ambulatory EDA data (Kleckner et al., 2018). The method evaluated signal quality in consecutive 2-s windows. First, the EDA level was checked to confirm that it was within the physiologically plausible range of 0.05–60 µS. Second, the rate of change was examined; windows exhibiting abrupt changes exceeding 10 µS s*^−^*^1^ were flagged as invalid, as such rapid fluctuations were typically caused by motion artifacts or sensor instability rather than true physiological responses. Only windows passing both criteria were retained for subsequent analyses. If we could not construct a matched control segment for a given arousal because of poor signal quality or excessive numbers of arousals or apneic events, we excluded the entire recording.

### 2.10 Arousal Labeling

The sleep arousal labels were not available in the dataset. Therefore, we detected arousals automatically using an open-source algorithm (Fernández-Varela et al., 2017) that has been independently validated and shown robust performance across diverse datasets (Alvarez-Estevez and Fernández-Varela, 2019). The algorithm operated on the PSG-derived C4–M1 EEG channel, the chin EMG signal, and the manually labeled sleep stage annotations. Within 3-second sliding windows, the algorithm computed alpha band power and beta band power as features. As an additional feature, EEG signal amplitude changes were also calculated. Candidate arousals were identified when these features exceeded predefined thresholds (alpha power *>* 0.25, beta power *>* 0.4, amplitude *>* 4).

For each candidate, EMG activity within and around the event was evaluated to refine event boundaries and estimate arousal duration. The algorithm then applied exclusion rules derived from the scoring guidelines (Berry et al., 2017), including the 3 s minimum arousal duration and the requirement of at least 10 s of preceding stable sleep. Events not meeting these criteria were discarded.

We implemented all signal processing in MATLAB R2021b (The MathWorks, Massachusetts, USA) and used R (Version 4.5.1) R Core Team (2025) with lme4 Bates et al. (2015), lmerTest Kuznetsova et al. (2017), and emmeans Lenth and Piaskowski (2026) packages for all statistical analyses.

## 3 RESULTS

From the initial cohort of 100 participants, we excluded one participant (ID 97) due to constant EDA values throughout the entire recording and 17 participants due to the diagnosed RLS. We also excluded two participants (ID 46 and ID 59) because their unusually high numbers of arousals and apneic events prevented the selection of a sufficient number of matched control segments.

The final dataset included 80 participants with a mean age 54.21 ± 16.29 (Table 2). Across these participants, we detected 4741 arousal events. After removing events overlapping with apnea, wake epochs, segments containing low-quality EDA, or segments with high activity in W-1, we retained 1583 arousal segments and an equal number of matched control segments. The REM sleep included 221 arousal and control segments and NREM sleep included 1362 arousal and control segments.

**Table 2.** Demographic, clinical, and dataset characteristics of the study population. Values are reported as mean ± SD.

| **Characteristic** | **Value** | **Description** |
| --- | --- | --- |
| **Number of participants** | 80 | Final cohort size |
| **Female (%)** | 42 (52.5%) | Sex distribution |
| **Age (years)** | 54.21 ± 16.29 | Age distribution |
| **BMI (kg/m<sup>2</sup>)** | 33.35 ± 25.59 | Body–Mass Index |
| **AHI (events/h)** | 10.71 ± 20.14 | Apnea–Hypopnea Index |
| **OAHI (events/h)** | 8.20 ± 17.85 | Obstructive AHI |
| **AI (events/h)** | 30.01 ± 23.40 | Arousal Index |
| **MAD (s)** | 7.13 ± 3.73 | Mean Arousal Duration |

The frequency analysis showed that band 8 was the transition from steep to gradual decline for both arousal-onset and reference windows (Figure 3). As bands 1–4 preceded this transition and exhibited higher mean power, we used them for extracting SLSymp. When averaged from 20 s before to 100 s after arousal onset across all participants, the mean SLSymp signal showed a clear post-onset increase, reflecting a characteristic sympathetic activation (Figure 4 (A)). The complementary signal calculated with the excluded frequency bands showed a similar overall rise in activation, but with a noisier trajectory and greater variability across participants than the dominant-band signal (Figure 4 (B)). This indicated that the dominant frequency bands captured the arousal-related sympathetic response more consistently and supported our choice of bands 1–4 for computing SLSymp.

**Figure 3.**
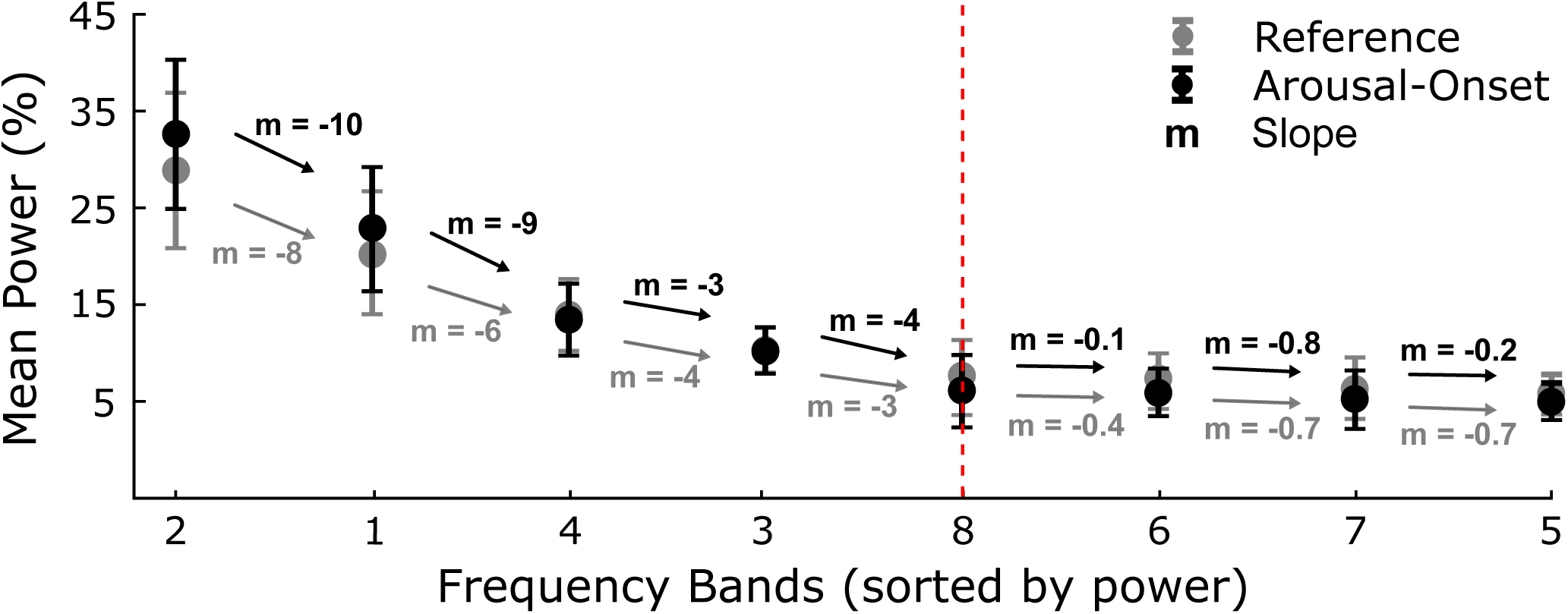
Sorted mean bandpower contributions with standard error bars across 1-min arousal-onset and reference windows. We sorted the extracted frequency bands Table 1) by mean power in descending order. Arrows indicate the slope (*m*) between consecutive ranked bands for the arousal-onset condition. The red dashed line marks the point of maximum slope ratio between consecutive slopes, indicating where the rate of power decline drops most sharply. We defined the bands ranked above this transition as dominant and used for SLSymp extraction.

**Figure 4.**
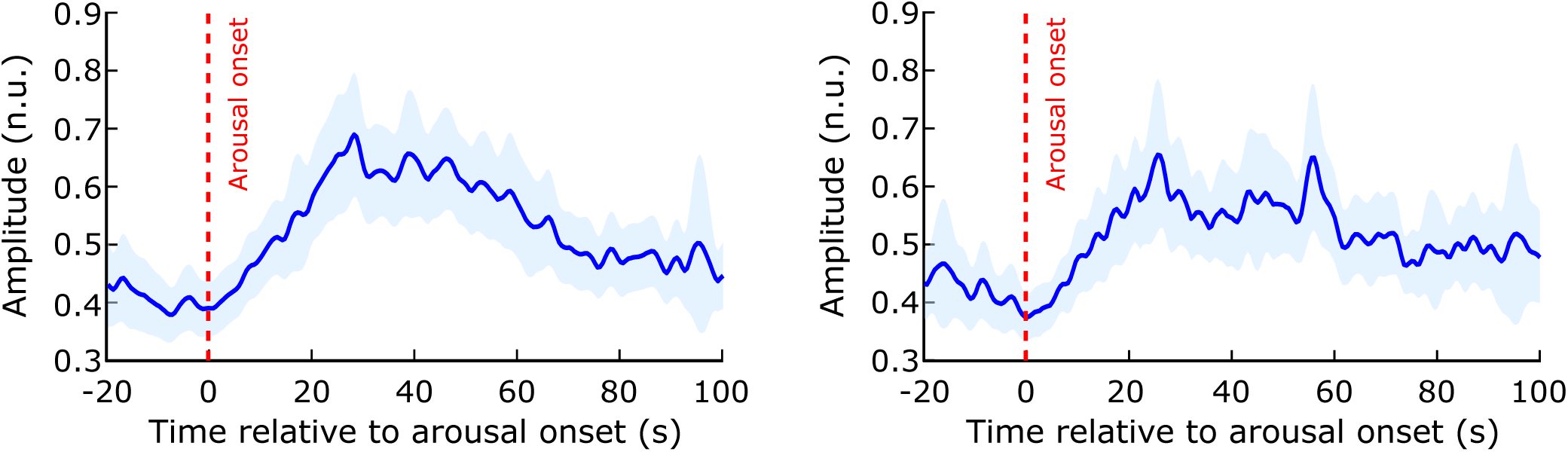
Temporal dynamics of sympathetic response, showing the time course from 20 s before to 100 s after sleep arousal onset. **(A)** Mean SLSymp amplitude across all arousals and participants. A clear post-onset rise is visible, with peak activation occurring approximately 30 s after arousal onset followed by a gradual return toward baseline. **(B)** Mean amplitude of the complementary signal, computed from the non-dominant frequency bands excluded from SLSymp. The rise and decay pattern in **(A)** shows less variability across participants than the complementary signal in **(B)**. The blue shaded regions represent the 95% confidence interval and the red vertical dashed line marks arousal onset.

Across all EDA-derived features, arousal segments exhibited approximately threefold larger rise and decay amplitudes than matched control segments, indicating stronger sympathetic activation during arousals (Figure 5). Feature values increased with arousal onset and remained elevated for up to approximately 40 s afterward. Window-to-window comparisons supported this pattern. All features showed significant increases from W-1 to W1, W2, W3, and W4 (*p <* 0.001), confirming a robust SLSymp response at arousal onset that persisted for several tens of seconds rather than returning immediately to baseline. This sustained profile contrasted with the HR response, which showed a sharp and significant increase during the arousal but returned to baseline immediately afterward (Figure 6).

**Figure 5.**
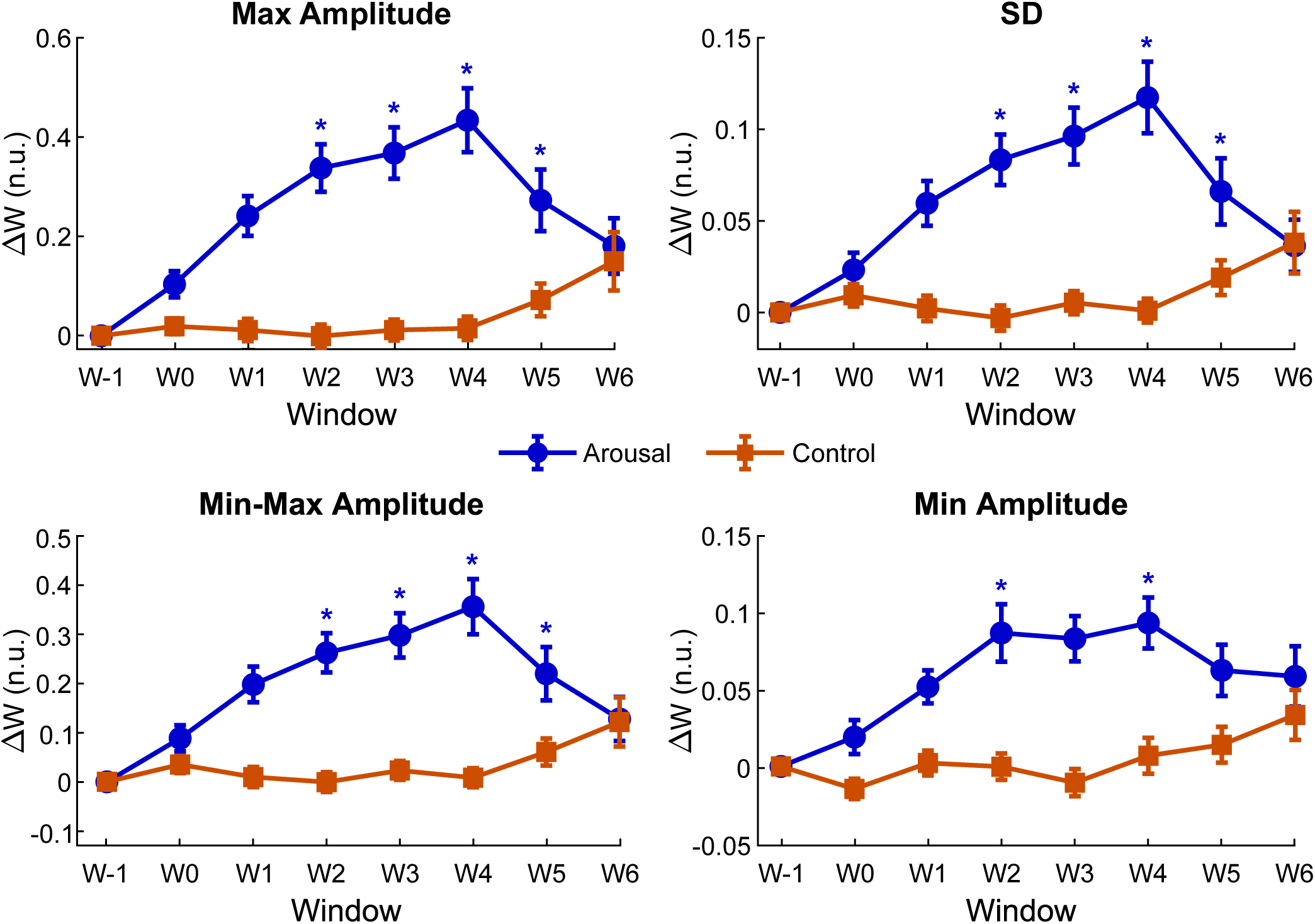
Baseline corrected trajectories of SLSymp features across arousal and control segments. Each panel shows the mean ΔW feature values with standard error bars across all arousal (blue) and matched control (orange) segments for four amplitude-based features: **(A)** maximum amplitude, **(B)** standard deviation, **(C)** min–max amplitude range, and **(D)** minimum amplitude. Mean feature values are plotted across seven temporal windows (W-1, W0, W1–W6). Across features, arousals are associated with a rise in amplitude-related metrics immediately after onset, followed by a gradual return toward baseline, whereas control segments remain comparatively stable. Asterisks mark significant increases (*p <* 0.001) in windows compared with W-1.

**Figure 6.**
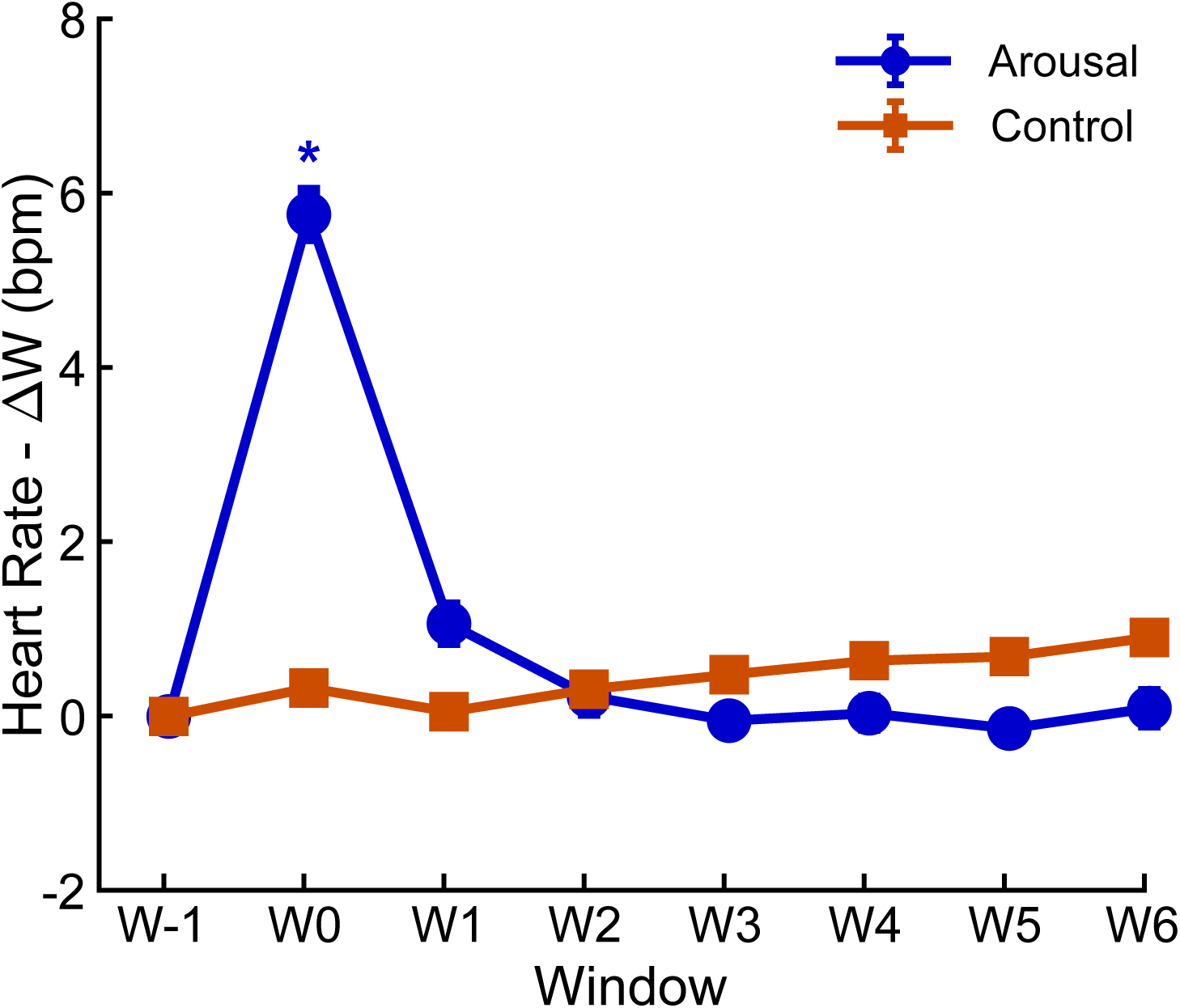
Baseline-corrected mean heart rate trajectories during arousal and control segments across seven temporal windows (W-1, W0, W1–W6). The mean heart rate changes from the pre-arousal baseline (W-1), with standard error bars are shown for arousal (blue) and matched control (orange) segments. Heart rate exhibits a sharp transient increase at W0, immediately following arousal onset, with rapid return to baseline. Control segments show a comparatively stable trajectory across all windows. Asterisk marks a significant increase (*p <* 0.001) in the arousal condition compared with W-1.

When analyzing the features by sleep stage, we selected the maximum amplitude feature as a representative measure of the SLSymp response. Arousal-related trajectories followed a broadly similar temporal pattern in REM and NREM sleep, with a rise after onset and a gradual decay (Figure 7). However, the two stages differed in their window-to-window significance. NREM showed significant increases from W-1 continuously across W1 through W4, whereas REM showed significance at W2 and W4 (*p <* 0.001). REM sleep also showed greater inter-individual variability, consistent with the higher and more heterogeneous sympathetic activity typically observed during this stage.

**Figure 7.**
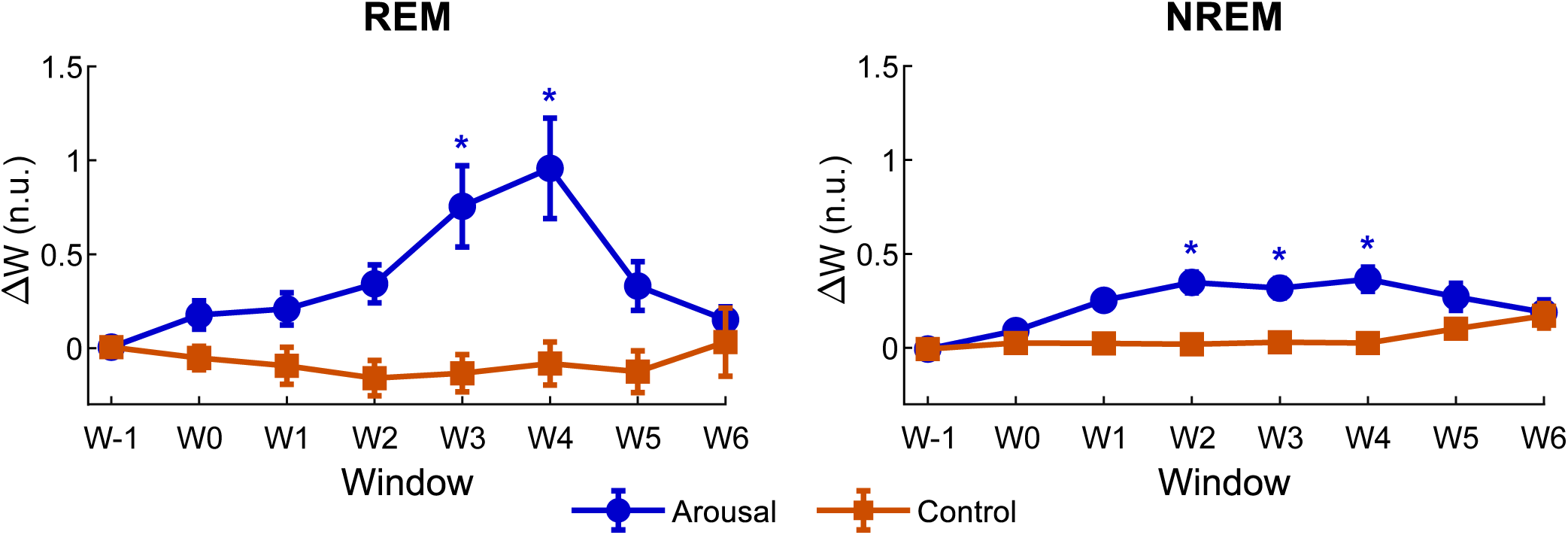
Baseline corrected SLSymp maximum amplitude feature trajectories during arousals for REM and NREM sleep. Each panel shows the mean ΔW feature values with standard error bars across seven temporal windows (W-1, W0, W1–W6), illustrated separately for **(A)** REM and **(B)** NREM sleep. Across both sleep stages, arousal onset produces a pronounced rise and gradual decay, with REM segments exhibiting greater variability. Asterisks mark significant increases (*p <* 0.001) in windows compared with W-1.

Because all features showed highly similar temporal patterns across segments (Figure 5) and there were consistent increases from W-1 to W1–W4, we reported the arousal duration analysis using a representative feature (max amplitude) to avoid redundancy. The window × duration interaction was significant (*F* (7, 12565) = 6.50, *p <* 0.001), indicating that longer arousals produced stronger EDA responses in the post-arousal period. Specifically, SLSymp features at W3 and W4 showed significant positive associations with arousal duration (*p <* 0.001), after Holm correction. Grouping arousals into three categories based on the 25th and 75th percentiles (4 s and 9 s, respectively) of the duration distribution confirmed this pattern. SLSymp trajectories diverged progressively with duration, with long arousals reaching the highest peak response (Figure 8).

**Figure 8.**
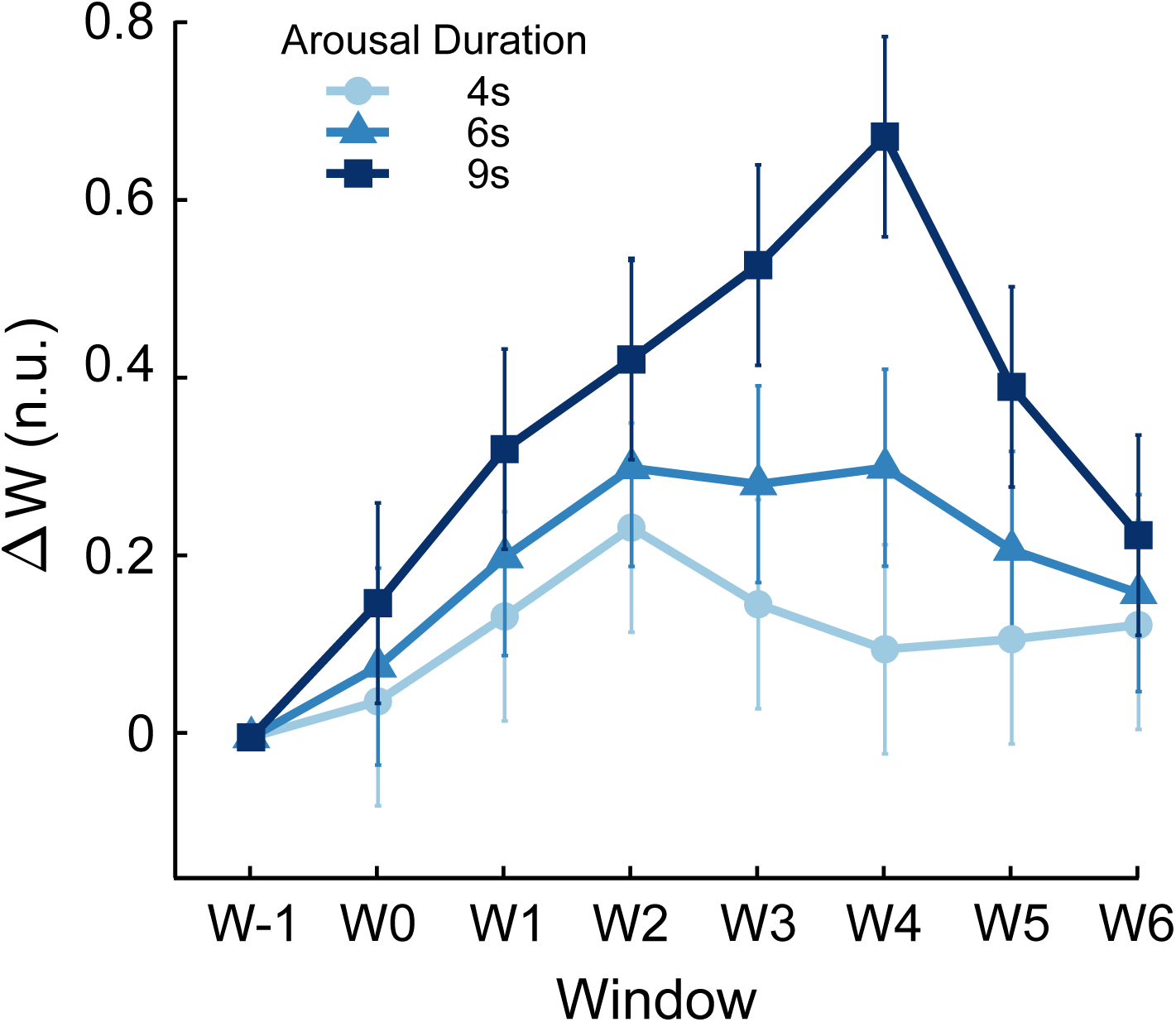
Baseline corrected SLSymp maximum amplitude feature trajectories across arousal windows for three arousal duration groups, defined using the 25th and 75th percentiles of the duration distribution (4 s and 9 s, respectively), with standard error bars. Longer arousals are associated with higher feature responses.

Across all peak-count criteria used to define storms, arousal segments contained a higher proportion of storm-positive epochs than matched control segments (Figure 9). This finding indicates that EDA storms are more likely to occur in temporal association with arousal events. Notably, the difference between arousal and control segments segments remained stable across thresholds, suggesting that this association does not depend on the specific storm definition.

**Figure 9.**
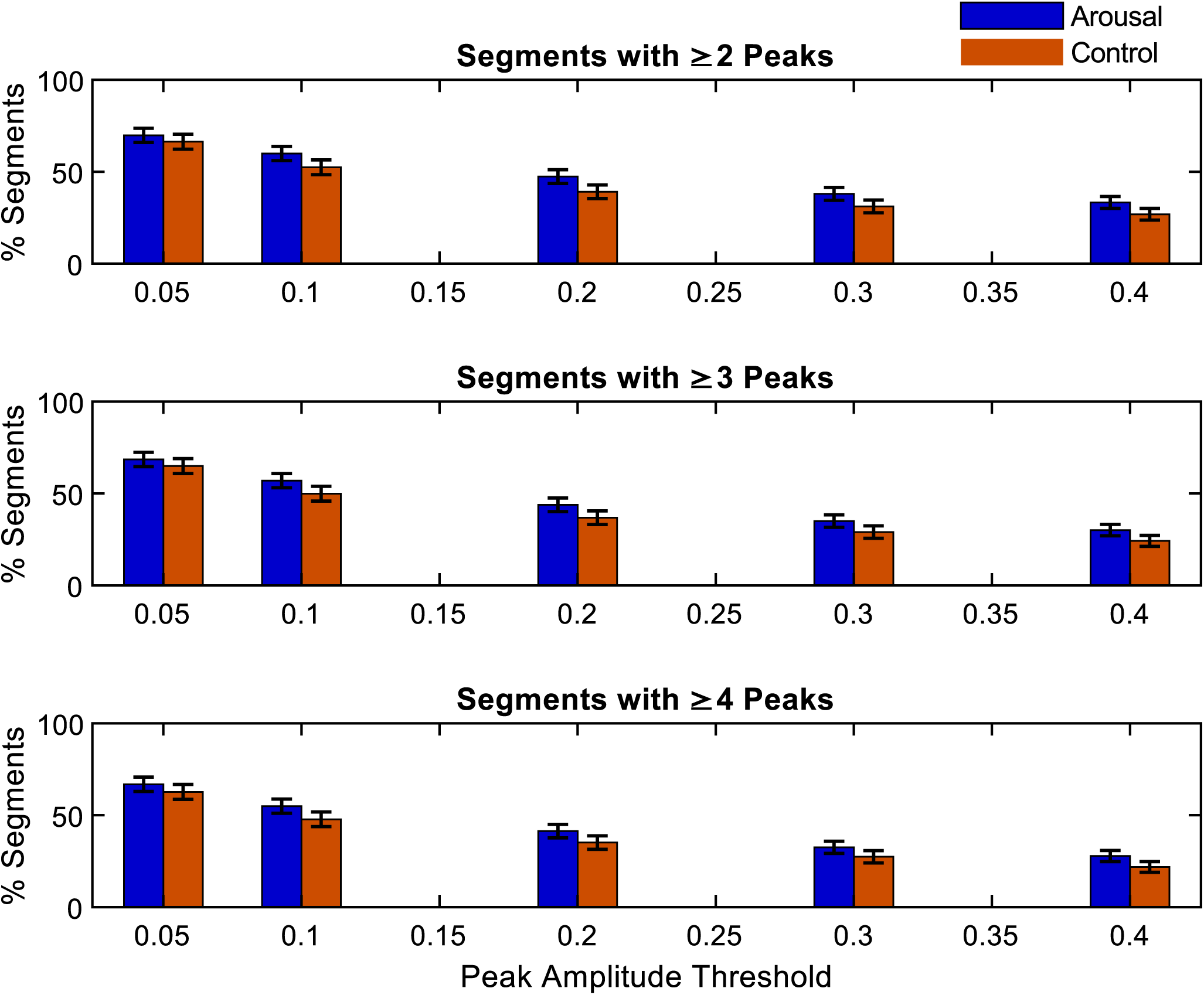
Percentage of segments containing ”storm” across varying peak amplitude thresholds. Each row corresponds to a different minimum peak-count criterion used to define a storm (top: at least two peaks, middle: at least three peaks, and bottom: at least four peaks). For each amplitude threshold on the x-axis, the y-axis shows the percentage of segments classified as storms. Arousal segments (blue) and matched control segments (orange) are displayed side by side to illustrate differences in storm occurrence across conditions.

The mean peak ACC amplitude of control segments was 0.02 g and the movement intensity was higher during arousal segments than during control segments across most windows (Figure 10). This was consistent with arousals commonly involving some degree of wrist movement. However, movement intensity varied substantially across arousals, and a large subset showed minimal activity throughout all windows, which was comparable to the levels observed in control segments.

**Figure 10.**
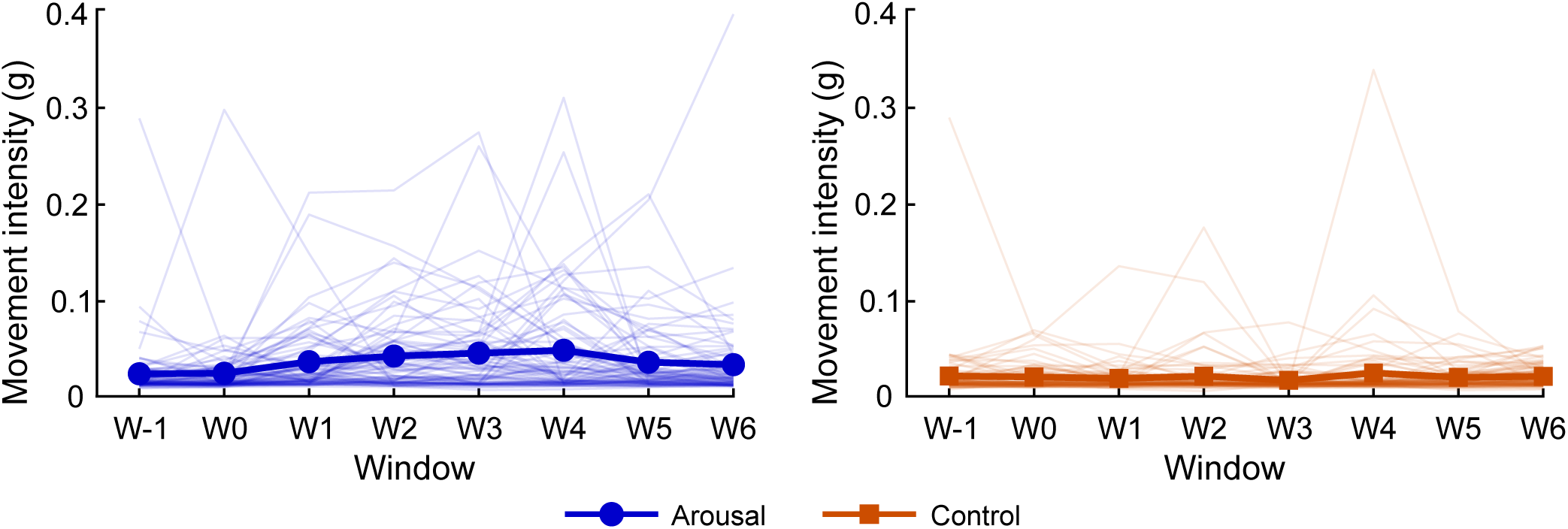
Movement intensity across all arousal and control segments. Thin lines show individual segment trajectories, and thick lines show the mean movement intensity across participants for arousal (blue) and control (orange) segments.

Arousals with low activity across all windows exhibited a rise-and-decay SLSymp trajectory consistent with the main event analysis, yet no window reached statistical significance (Figure 11 (I)). For arousals grouped by the window of peak movement intensity, the SLSymp peak closely tracked the timing of movement: in each group from W0 through W6, the window of high activity coincided with a significant SLSymp increase relative to W-1, after Holm correction (*p <* 0.01, Figure 11 (B)–(H)). Arousals with high activity in W-1 were an exception, showing a below-baseline SLSymp trajectory rather than an increase (Figure 11 (A)).

**Figure 11.**
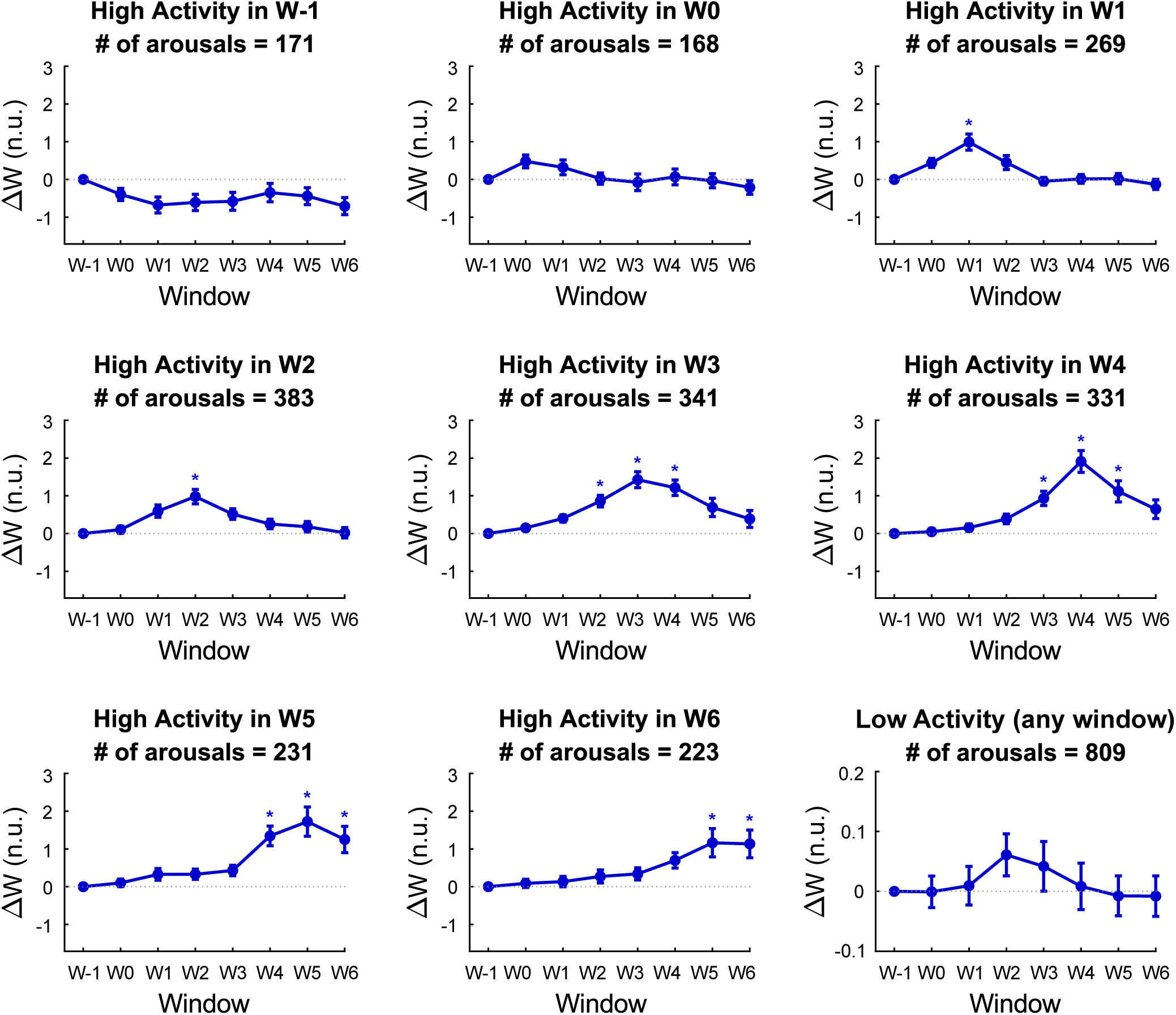
SLSymp maximum amplitude feature trajectories for arousals grouped by the window of high activity. Panels **(A) – (H)** show arousals classified as high activity in W-1 through W6, respectively, grouped by the specific window in which their movement intensity exceeded the activity threshold. Panel **(I)** shows arousals with low activity across all windows. Error bars represent the standard error of the mean and event counts per group are indicated in each panel. The number of arousal events in each group is given in their respective panels. Asterisks mark significant changes (*p <* 0.01) in windows compared with W-1.

## 4 DISCUSSION

We conducted a multimodal analysis of sudomotor, cardiac, and motor dynamics during sleep arousals, which showed that arousal-related sympathetic activation is more sustained and distinct from cardiac and motor responses. SLSymp features remained elevated for approximately 40 s after arousal termination, well beyond the rapid, transient heart rate response and independent of physical movement.

In this study, we showed that sleep arousals are accompanied by marked increases in SLSymp features, with elevated sympathetic response persisting for approximately 40 s after arousal termination. We systematically characterized the temporal dynamics of electrodermal activity around sleep arousals using a decomposition approach tailored for sleep. By comparing arousal segments with matched control segments, we demonstrated that arousals were associated with a distinct and sustained sympathetic response beyond the background fluctuations observed during stable sleep. In contrast, heart rate exhibited a rapid, transient response that was largely confined to the arousal itself, highlighting the complementary physiological information provided by EDA. In addition, arousal segments exhibited a higher prevalence of EDA storms than control segments, further supporting the close temporal association between arousals and transient bursts of sympathetic activation. Although wrist movement was associated with enhanced SLSymp responses, elevated sympathetic response was also observed during the low activity, indicating that movement alone does not account for the observed EDA dynamics.

The amplitude-based feature analysis revealed consistent increases across all metrics immediately after arousal onset. All the examined measures showed significant elevations in the W2 to W4 windows relative to the W-1 and W0 windows. These temporal patterns suggested a characteristic rise-and-decay response, indicating that the effect of arousal-related sympathetic activation persist up to 40 s before returning toward baseline. The sustained increases likely reflected both continued sympathetic sudomotor activity after arousal termination and the slow recovery of sweat gland function, where secreted sweat persists before evaporating or being reabsorbed.

This sustained rise-and-decay pattern contrasted markedly with the heart rate response to arousals, which increased sharply during the arousal itself but returned to baseline immediately afterward. This dissociation suggested that EDA and HR capture distinct aspects of the arousal response. HR reflected a rapid, transient autonomic reflex tightly coupled to the arousal event, whereas EDA reflected a slower, more sustained sympathetic engagement that outlasted the arousal itself. This distinction reinforced the value of EDA as a complementary marker capturing autonomic dynamics not evident from cardiac measures alone.

Arousal duration further modulated the post-arousal EDA response in a graded, continuous fashion. Longer arousals produced systematically stronger EDA responses across the post-arousal period, specifically at W3 and W4. These results suggest that arousal duration does not simply determine whether a sympathetic response occurs, but scales the magnitude and temporal persistence of the sudomotor activation. Longer arousals may sustain sympathetic outflow for a greater duration, resulting in more secreted sweat that remains detectable in the mid-to-late post-arousal windows. These findings extend prior observations that arousal duration influences the autonomic response magnitude (Azarbarzin et al., 2014), and suggest that this influence is graded and window-specific.

Feature trajectories were similar between REM and NREM sleep, but REM segments exhibited noticeably higher variability. This larger variance may reflect the intrinsically fluctuating autonomic environment of REM sleep, where sympathetic activity is known to be elevated. It is also important to note that the number of REM segments was substantially smaller than that of NREM, which may have contributed to the increased variability through reduced averaging.

Our analysis showed that arousals markedly increased the likelihood of EDA storms. Prior work demonstrated that storms occur naturally throughout sleep as part of baseline sympathetic activity (Sano et al., 2014). We observed this same phenomenon in our data: both arousal and control segments exhibited storms, confirming that spontaneous peak bursts are an expected component of normal sleep physiology. Our study also showed that arousal segments consistently included a higher proportion of storms. Importantly, this increase did not coincide with a higher number of peaks within storms compared to control segments. Thus, arousals appear to elevate the probability that a storm occurs rather than altering the storm structure itself.

When interpreting storm dynamics together with the statistical SLSymp features, a coherent pattern emerges. Arousal onset produced robust increases in amplitude-based features but did not yield additional peaks within storms. This indicates that the sympathetic response to arousals manifests primarily as enhanced magnitude rather than increased frequency of electrodermal events. One possible physiological explanation is the transient synchronization of sweat gland activity during arousals. Increased sympathetic outflow during arousal may recruit additional glands or enhance temporal coordination, producing larger, more spatially coherent skin conductance responses without requiring additional discrete sympathetic bursts. As a result, peaks that were too small to detect during stable sleep become detectable during arousal, increasing storm likelihood without proportionally increasing peak counts. This represents a shift in the amplitude distribution of sympathetic activity rather than a change in discharge frequency, consistent with arousal-induced reorganization of autonomic output patterns.

Movement did not appear to be the primary driver of the observed SLSymp responses. Arousals showed a genuine SLSymp response, even in the absence of high wrist activity, consistent with our main findings. Although this trajectory did not reach statistical significance, the visible elevation across post-arousal windows suggested that a genuine sympathetic response was present. The statistical results were likely due to the smaller sample size of low-movement events reducing statistical power. Movement was, on average, higher during arousals than during stable sleep, in line with arousals commonly involving some degree of physical activity. However, a substantial subset of arousals showed minimal movement, comparable to control-segment levels, and these low-movement arousals still exhibited the characteristic rise-and-decay SLSymp trajectory observed in the main analysis. This indicated that the sympathetic response to arousal is not contingent on movement. When high movement activity was present, it was associated with a larger SLSymp response, suggesting that movement amplified rather than caused the SLSymp changes observed around arousals.

While this study provides a robust characterization of arousal-related sympathetic dynamics, several factors define its scope. First, the analysis focused on discrete arousal events without accounting for the potential modulatory effects of concurrent sleep microstructures, such as K-complexes, spindles, or slow-wave activity, which are known to influence autonomic tone (de Zambotti et al., 2018). Second, to maintain statistical power across the cohort, sleep stages were analyzed as REM and NREM; future work should explore the finer granularity of N1–N3 substages and the tonic-phasic dichotomy of REM sleep. Additionally, although the automated arousal detection algorithm has been extensively validated, the lack of manual annotations in this specific dataset introduces a potential for minor detection bias, which would likely add noise rather than systematically bias the arousal-versus-control comparisons. Finally, while this study established a general population baseline, the modulatory role of sex hormones on sudomotor responses was not evaluated and remains a critical area for subsequent investigation.

## 5 CONCLUSION

This work provided a comprehensive characterization of sympathetic response during sleep arousals using SLSymp, an EDA decomposition tailored for sleep. By jointly examining amplitude-based SLSymp features, temporal trajectories, arousal-duration effects, sleep-stage effects, and storm dynamics, we showed that arousals elicit a robust, sustained sympathetic response that extends well beyond arousal termination. Our results revealed a consistent rise-and-decay pattern across features, shaped primarily by stronger sudomotor responses rather than more frequent discrete events. This temporal profile dissociated from heart rate and indicated that EDA captured a slower and more sustained facet of sympathetic response, which was not reflected in the rapid, transient cardiac response to arousal. The persistence of this sustained response even under minimal movement further indicated that it reflected sympathetic activation rather than motion artifacts, though movement amplified the response magnitude when present. These findings highlight the potential of wearable EDA as a complementary modality for unobtrusive assessment of sympathetic activation during sleep and provide a physiological foundation for future wearable sleep monitoring applications.

## Data Availability

Data used in this study is openly available online at: https://doi.org/10.13026/dztc-dv77

## AUTHOR CONTRIBUTIONS

TCG: Conceptualization, Formal analysis, Funding acquisition, Investigation, Methodology, Software, Visualization, Writing – original draft, Writing – review & editing. HPQ: Conceptualization, Methodology, Software, Writing – review & editing. YK: Conceptualization, Methodology, Software, Writing – review & editing. CJW: Conceptualization, Writing – review & editing. KC: Conceptualization, Supervision, Writing – review & editing. WK: Conceptualization, Funding acquisition, Methodology, Supervision, Writing – review & editing.

## ACKNOWLEDGMENTS

We thank the members of the Chon Lab, the Posada-Quintero Lab, and the SleepLoop consortium for many insightful discussions. We also thank the creators of the open-access dataset DREAMT and Physionet for making the data publicly accessible.

## GENERATIVE AI STATEMENT

The author(s) declared that generative AI was used in the creation of this manuscript. The authors used Claude (Anthropic) to support English grammar refinement and improve clarity. The authors reviewed and edited all AI-assisted content and take full responsibility for the accuracy and integrity of the published work.

## DATA AVAILABILITY STATEMENT

The open-source dataset analyzed for this study can be found in the Physionet platform (https://www.physionet.org/content/dreamt). The automated sleep arousal detection algorithm is openly available on GitHub (https://github.com/bigsasi/arousals-detection). EDA quality assessment algorithm is also openly available on GitHub (https://github.com/iankleckner/EDAQA). To support reproducibility, we share our code for participant selection and ECG and ACC pre-processing on GitHub (https://github.com/tugcecanbaz/sleep-arousal-EDA).

